# Does Heat Thermotherapy Improve Cardiovascular and Cardiometabolic Health? A Systematic Review and Narrative Synthesis of the Literature

**DOI:** 10.1101/2020.05.19.20106666

**Authors:** Ben S. Price, Samuel J. E. Lucas, Rachel E. Gifford, Nigel T. Cable, Rebekah A. I. Lucas

## Abstract

**Objective:** To assess the efficacy of heat thermotherapy to improve cardiovascular and cardiometabolic health, and to compare potential heat thermotherapy moderating factors.

**Design:** Systematic review and narrative synthesis.

**Data Sources:** Electronic databases (MEDLINE, EMBASE and Web of Science) were searched up to 6th February 2020.

**Eligibility criteria for selecting studies:** Journal articles with adult participants, a controlled trial study design, and a passive heating stimulus with a cardiovascular or cardiometabolic health outcome were included.

**Results:** From 1036 articles, 39 articles met the inclusion criteria. Heat thermotherapy was delivered acutely (one bout; n=19), short term (2-15 bouts; n=6) and chronically (>15 bouts; n=14), via either hot water immersion (n=26; water temperature 38-43 °C) or heated air exposure (n=11; air temperature 31-90 °C), or water perfused suit/handheld device (n=2). Heat exposure ranged from 10 to 240 minutes. Cardiovascular and cardiometabolic measurement techniques varied across studies, alongside participant age (≤35 years, n=376; >35 ≤60 years, n=335 and >60 years, n=350) and health status. 21/27 studies measuring cardiovascular outcomes reported positive health benefits, including increased flow-mediated dilation (1.3-5.3%) and lower systolic blood pressure (4-16 mm Hg). 15/22 studies measuring cardiometabolic outcomes reported positive health benefits, including reduced postprandial glucose (−0.89 to −0.5 mmol.L^-1^) and C-reactive protein concentrations (−0.9 to −10.94 mg.L^-1^).

**Conclusion:** Overall, 29/39 studies demonstrated significant positive cardiovascular or cardiometabolic health benefits from heat thermotherapy across various population demographics, despite varied study design and modality. Heat thermotherapy is an efficacious treatment for cardiovascular disease alongside pharmaceutical and exercise-based interventions.

## INTRODUCTION

Cardiovascular disease (CVD) is the leading non-communicable cause of death worldwide.^1^ Main risk factors include hypertension, obesity and physical inactivity, all of which are associated with a sedentary lifestyle.^2^ Typical approaches to prevent and treat CVD include pharmaceutical and exercise interventions (e.g. 150 min/week of moderate-intensity or 75 min/week of vigorous-intensity aerobic exercise).^3^ Pharmaceutical interventions, however, are costly and have a low adherence rate. For example, over 50% of patients after 12 months stop taking antihypertensive medication.^4^ Similarly, despite its efficacy to reduce CVD risk,^2^ exercise also has low adherence rates due to factors such as time constraints, cost of participation (i.e. gym memberships) and confidence.^5^ Therefore, there is a demand for alternative interventions that are cost-effective, pragmatic and have a potential for higher adherence rates than traditional intervention strategies.

Heat Thermotherapy (HT) is the application of a passive (non-exercising) heating stimulus that increases core body temperature and results in a beneficial health outcome. Recent literature has shown both positive cardiometabolic benefits^6^ and positive cardiovascular benefits of HT.^7^ This includes reductions in blood pressure, inflammatory markers and improved endothelial functioning, which are associated with a reduction in cardiovascular-related mortality.^2 8^ These cardiovascular and cardiometabolic improvements have been seen in numerous populations from young sedentary, healthy cohorts to chronic heart failure patients. Furthermore, HT adherence rates are higher than exercise interventions.^9^ Therefore, initial evidence indicates that HT is potentially a pragmatic and easily accessible method to help prevent non-communicable disease progression in a variety of population groups.

Despite the promise of HT to improve both cardiovascular and cardiometabolic health, a systematic review of the literature to formally assess this conjecture has not been conducted to date. Moreover, the minimal and optimal conditions for HT to elicit beneficial cardiovascular and cardiometabolic health benefits are yet to be determined. Recent reviews have aimed to analyse the mechanisms for cardiovascular and cardiometabolic benefits during HT or to show positive associations between regular HT sessions and reduced allcause mortality rates.^10-12^ Despite the benefits of identifying the physiological mechanisms underpinning HT adaptations, these reviews have not directly analysed the methodology and subsequent results of HT. Furthermore, they include studies that focus on associative data, which may show the potential positive effects of HT but cannot establish causational relationships. Therefore, a systematic review is needed to assess whether HT improves cardiovascular and cardiometabolic health outcomes, including papers with causative study designs.

This systematic review and narrative synthesis aimed to investigate the balance of evidence for the efficacy of passive heat therapy in improving cardiovascular function and cardiometabolic health. Specifically, this review systematically reviewed causational study design literature and quantified different intervention conditions and subsequent cardiovascular and cardiometabolic responses. The narrative synthesis focused on potential key moderating factors such as the relationship between core body temperature change and subsequent cardiovascular and cardiometabolic outcomes. Thus, the combination of the systematic review and narrative synthesis will provide comprehensive evidence on whether HT can improve cardiovascular and cardiometabolic health. It will also, where possible, indicate the most promising methods to elicit positive cardiovascular and cardiometabolic health outcomes through HT.

## METHODS

### Overview

For this systematic review the PICO process, PRISMA guidelines were strictly adhered to including the development of a PRISMA flowchart.^13-15^ The narrative synthesis was conducted according to established guidelines to gather heterogeneous evidence with multiple outcomes in a transparent, non-bias, systematic way.^16^ The narrative synthesis was conducted in four stages: 1. developing a theory; 2. developing a preliminary synthesis; 3. exploring relationships, and 4. assessing the robustness of the synthesis.

### Study Selection Process

Studies were chosen using the patient, intervention, comparison, outcome (PICO^13^) process (*see online supplementary table 1*). The PICO was designed to ensure a broad array of HT studies with cardiovascular or cardiometabolic measurements were included. The criteria for participants were human adult (≥18 years old) studies. The population section included a broad range of terms to ensure both healthy participants and participants in cardiovascular and cardiometabolic preclinical or disease states were included. In the intervention section, the criteria were designed to capture different HT interventions that used passive heating to increase body temperature. The comparison section was designed to include journal articles in English with causational study design. The outcome section was designed to cover a range of cardiovascular and cardiometabolic measurements.

The study selection process is outlined in *Figure 1*. Two reviewers (BP, RG) screened the abstracts and titles of the 1036 citations retrieved in the database and manual searches with any disagreement concluded by a third reviewer (RL). From the 1036 abstracts reviewed, 119 papers were selected for the second screening process. This involved the reading of the full text of 119 papers and assessing their content according to the inclusion and exclusion criteria outlined above, with the addition that studies were also excluded if the full text was not available. Authors were contacted via email to retrieve any missing data. From the 119 full texts reviewed, 39 papers were selected for full data extraction and inclusion in the narrative synthesis.

**Figure 1.**
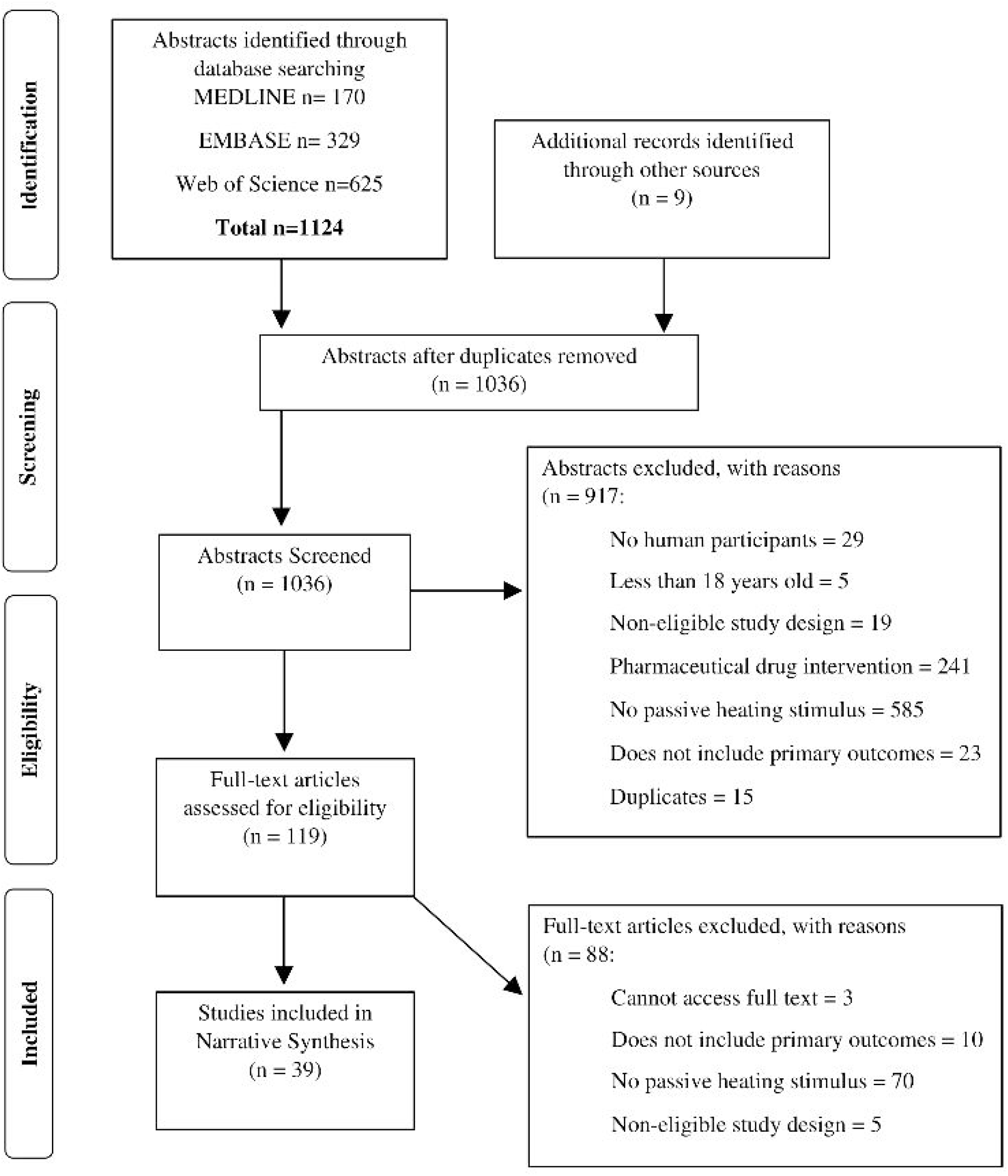

### Information Sources and Search Strategy

Bibliographic databases, MEDLINE, EMBASE and Web of Science, were formally searched as the primary means of identifying relevant texts (*see online supplementary table 2*). An additional manual literature search was conducted, and nine papers were added to the initial screening phase. Databases and manual searches included texts from the earliest start date to the 6^th^ February 2020. In total 1036 papers were retrieved. Search results were extracted to EndNote (Clarivate Analytics, Philadelphia, Pennsylvania, USA) and duplicates were removed before continuing the screening process.

### Narrative Synthesis

#### Developing a Theory

A theory was developed to display the reasons and potential moderating variables for the efficacy of HT to improve cardiovascular and cardiometabolic health (*Figure 2*). This theory was then evaluated in the exploring relationships section of the narrative synthesis.

**Figure 2.**
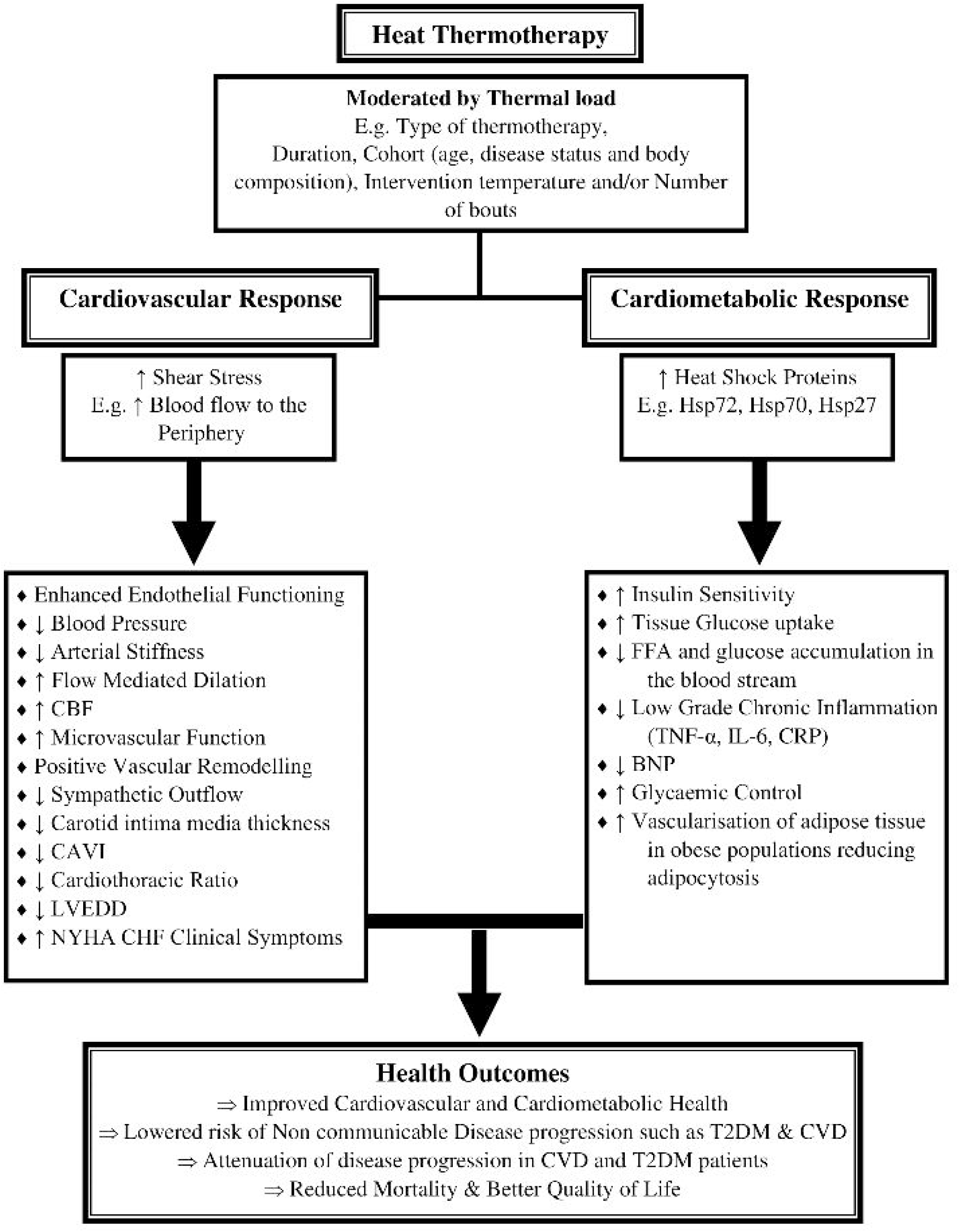

#### Preliminary Synthesis

A systematic search of the literature was conducted to investigate the HT and the positive cardiovascular/cardiometabolic health outcomes theory. The focus of the search was to locate literature of causational design that has measured cardiovascular or cardiometabolic health outcomes with an HT intervention. The narrative synthesis was created using the data extracted from 39 studies, which had a range of participants, study designs, intervention designs, body temperature changes and cardiovascular and cardiometabolic outcomes. The preliminary synthesis was completed in two stages. In stage one, the data from all included 39 studies were assessed by a common format (core body temperature change) and summarised to highlight the overall findings in terms of the number of participants alongside cardiovascular and cardiometabolic outcomes. In stage two, studies were grouped by different factors such as intervention type (bath vs sauna), duration of the stimulus and the number of bouts (acute vs short term vs chronic studies). Data considered to be of interest (e.g. average core body temperature change and cardiovascular outcomes) was tabulated to help identify patterns in the data.

#### Exploring Relationships

This stage investigated the relationships between HT conditions and cardiovascular/cardiometabolic outcomes within and between studies. Conceptual mapping and triangulation techniques were used to consider what information from the tables and figures was important enough to be included within the narrative synthesis. The focus of the techniques was to derive the moderating variables that may influence HT in terms of its efficacy to induce cardiovascular and cardiometabolic health benefits.

#### Assessing the Robustness of the Synthesis

The robustness of the narrative synthesis was assessed in two sections. Firstly, the amount and quality of the evidence in the included studies were assessed and secondly, the methods used to create the narrative synthesis was assessed. To address the first section, a Risk of Bias Assessment was conducted using COCHRANE Guidelines.^17^ This assessment determined the risk of type one or type two error by assessing five domains, specifically: potential bias arising from the randomisation process; potential bias due to deviations from intended interventions; potential bias due to missing outcome data; potential bias in the measurement of the outcome, and potential bias in the selection in the reported result (*see online supplementary table 3 for more details*). Two reviewers completed the risk of bias (BP and RL) with a third reviewer (SL) consulted to resolve any discrepancies. In section two, the methods in the narrative synthesis were critically evaluated, including any assumptions about the data reported and/or any methodological limitations, to give an overall assessment of the robustness of the synthesis.

## RESULTS

### Development of a Theory

The theoretical model was based on recent review papers in the literature^11 18^ and is illustrated in *Figure 2*. HT, as derived from the theory, increases core temperature, which drives cardiometabolic and cardiovascular responses. These cardiovascular responses subsequently result in health improvements (e.g. reduced blood pressure and inflammation) and reduce the risk of chronic diseases or attenuate disease progression^18^. Such health benefits could be moderated by HT methodology parameters, such as the duration or number of bouts the participant is exposed to.

### Preliminary Synthesis

Within the 39 included studies, 1061 participants completed one to over 127 separate thermotherapy bouts within the same intervention protocol. The HT modality characteristics are reported in *Table* 1. Heat thermotherapy was delivered acutely (one bout; n=19), short term (2-15 bouts; n=6) and chronically (>15 bouts; n=14), via either hot water immersion (n=26; water temperature 38-43 °C) or heated air exposure (n=11; air temperature 31-90 °C), or a water perfused suit/handheld device (n=2). Participant’s age for included studies was: ≤35 years, n=376; >35 ≤60 years, n=335 and >60 years, n=350. Participants health status was: Healthy (no risk factors or health conditions; n = 328), Cardiovascular risk factor (≥1) including being overweight/obese (n= 666) and congestive heart failure (any class; n = 67)).

**Table 1.** HT Method Characteristics

| **Descriptive Factor**<br>(Initial<br>Thermotherapy<br>conditions) | **Number**<br>**of**<br>**Papers** | **Mean**<br>**Duration**<br>**(minutes)** | **Duration**<br>**Range**<br>**(minutes)** | **Mean Core**<br>**Body**<br>**Temperature**<br>**Change (°C)** | **Core Body**<br>**Temperature**<br>**Change Range**<br>**(°C)** |
| --- | --- | --- | --- | --- | --- |
| <u>Hot Water Immersion</u> |  |  |  |  |  |
| Temperate water<br>temperature ($\leq 38$<br>°C) | 3 | 27 | 20-30 | Not Reported | Not Reported |
| Warm water<br>temperature ($> 38$;<br>$\leq 39.9$ °C) | 4 | 53 | 10-120 | 1.6 | 1.5-1.6 |
| Hot water<br>temperature ($> 39.9$<br>°C) | 14 | 44 | 10-90 | 1.5 | 0.6-1.8 |
| <u>Heated Air</u> |  |  |  |  |  |
| Moderately high air<br>temperature, low<br>humidity ($\leq 63$ °C;<br>$\leq 30\%$ RH) | 4 | 39 | 15-110 | 0.6 | 0.6-0.6 |
| Moderately high air<br>temperature, high<br>humidity ($\leq 63$ °C;<br>$> 30\%$ RH) | 1 | 240 | 240-240 | 0.8 | 0.8-0.8 |
| High air<br>temperature, low<br>humidity ($> 63$ °C;<br>$\leq 30\%$ RH) | 5 | 32 | 25-45 | 1.7 | 0.8-2.7 |
| High air<br>temperature, high<br>humidity ($> 63$ °C;<br>$> 30\%$ RH) | 1 | 16 | 16-16 | Not Reported | Not Reported |

*Note.* Eight papers were excluded from the table as they heated a single limb, feet only or used a water perfused suit. For HWI papers: core temperature was not reported in 3/3 of the temperate water ( $\leq 38$ °C) papers, 2/4 warm water ( $> 38$ ; $\leq 39.9$ °C) papers and 5/14 of the hot water ( $> 39.9$ °C) papers. For heated air papers, core temperature was not reported in 3/4 of the moderately high temperature, low humidity ( $\leq 63$ °C; $\leq 30\%$ RH) papers, 1/5 high temperature, low humidity papers ( $> 63$ °C; $\leq 30\%$ RH) and 1/1 of the high temperature, high humidity ( $> 63$ °C; $> 30\%$ RH) papers. Overall, 15/39 heated air and HWI papers included in the table did not report core temperature change.

The paper summaries for all 39 included studies outlining key details regarding environmental conditions, duration, and number of bouts during the experimental trial are shown in *Table 2*. Core body temperature was measured in 21/39 of the included studies. Studies that did report core body temperature used a range of measurement sites including rectal, oral, tympanic, and auxiliary. The most frequently reported cardiovascular measures was blood pressure (MAP, SBP and DBP respectively). The most frequently reported cardiometabolic measures was glucose concentration (fasting, during and postprandial collectively). Overall, 74% of studies reported either a positive cardiovascular or cardiometabolic health benefit because of an HT intervention (*Table 3*). Of those papers, the breakdown of potential moderating thermotherapy intervention parameters and the cardiovascular/cardiometabolic outcomes are reported in *Figure 3*.

**Table 2.**
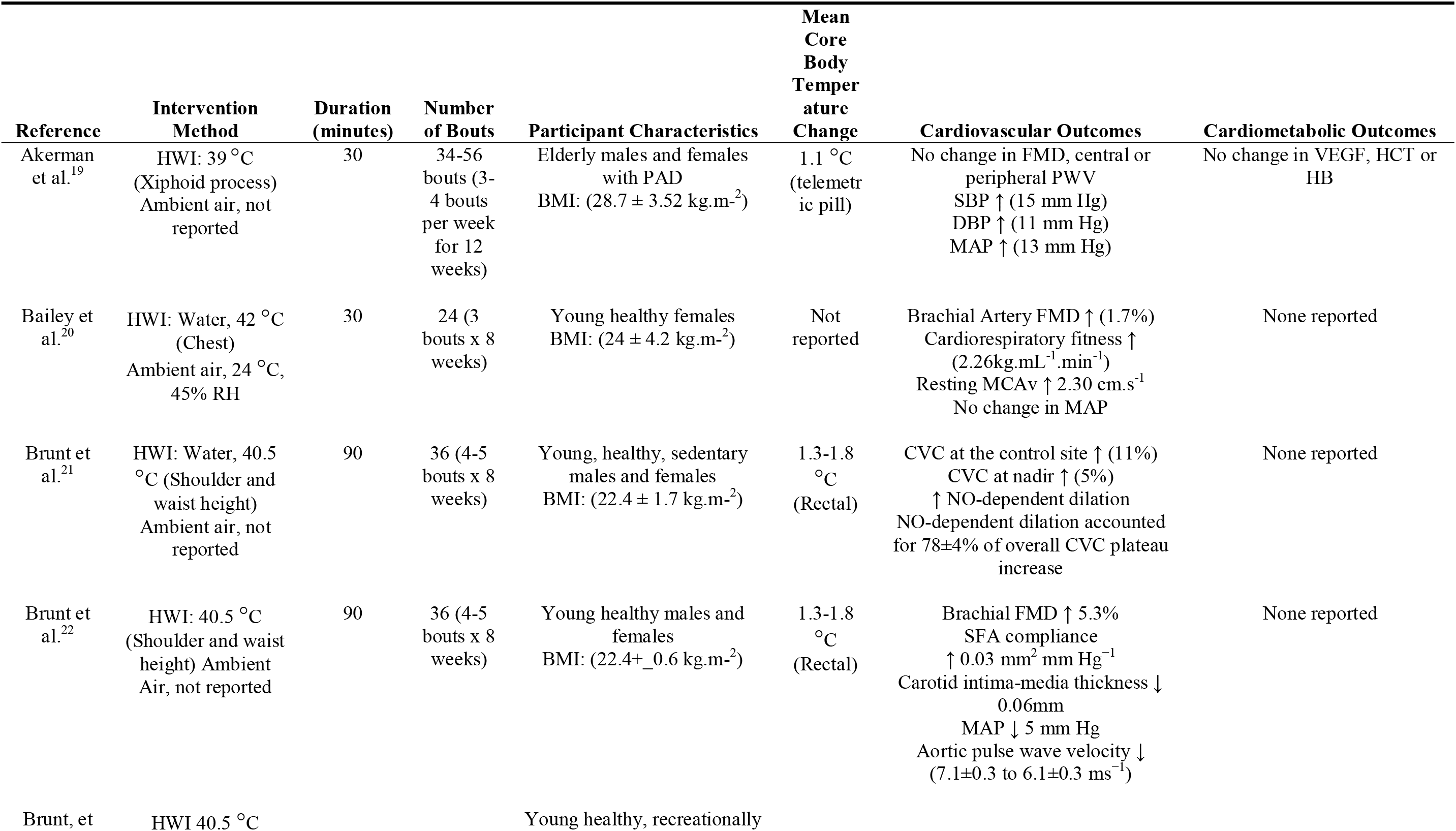
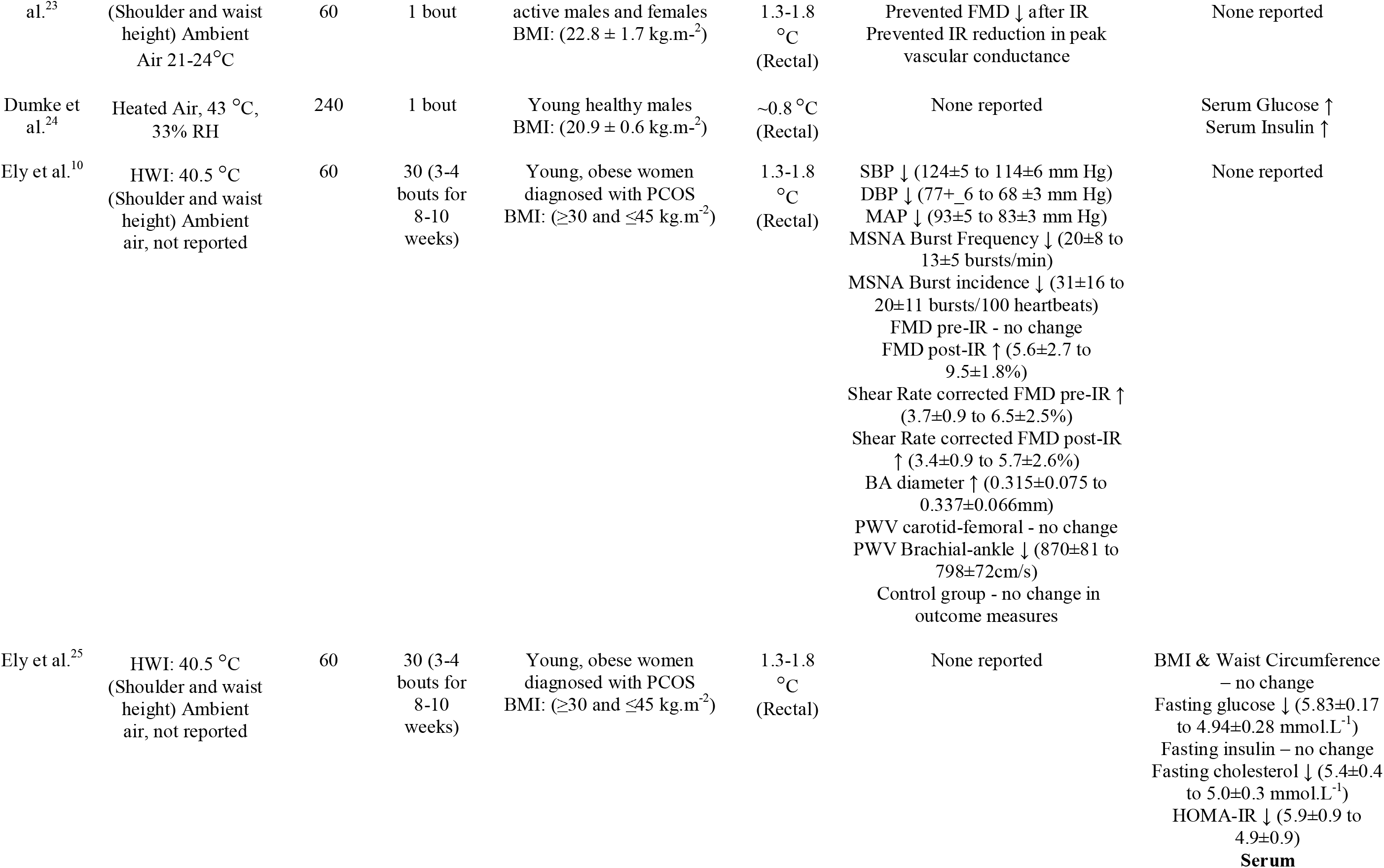
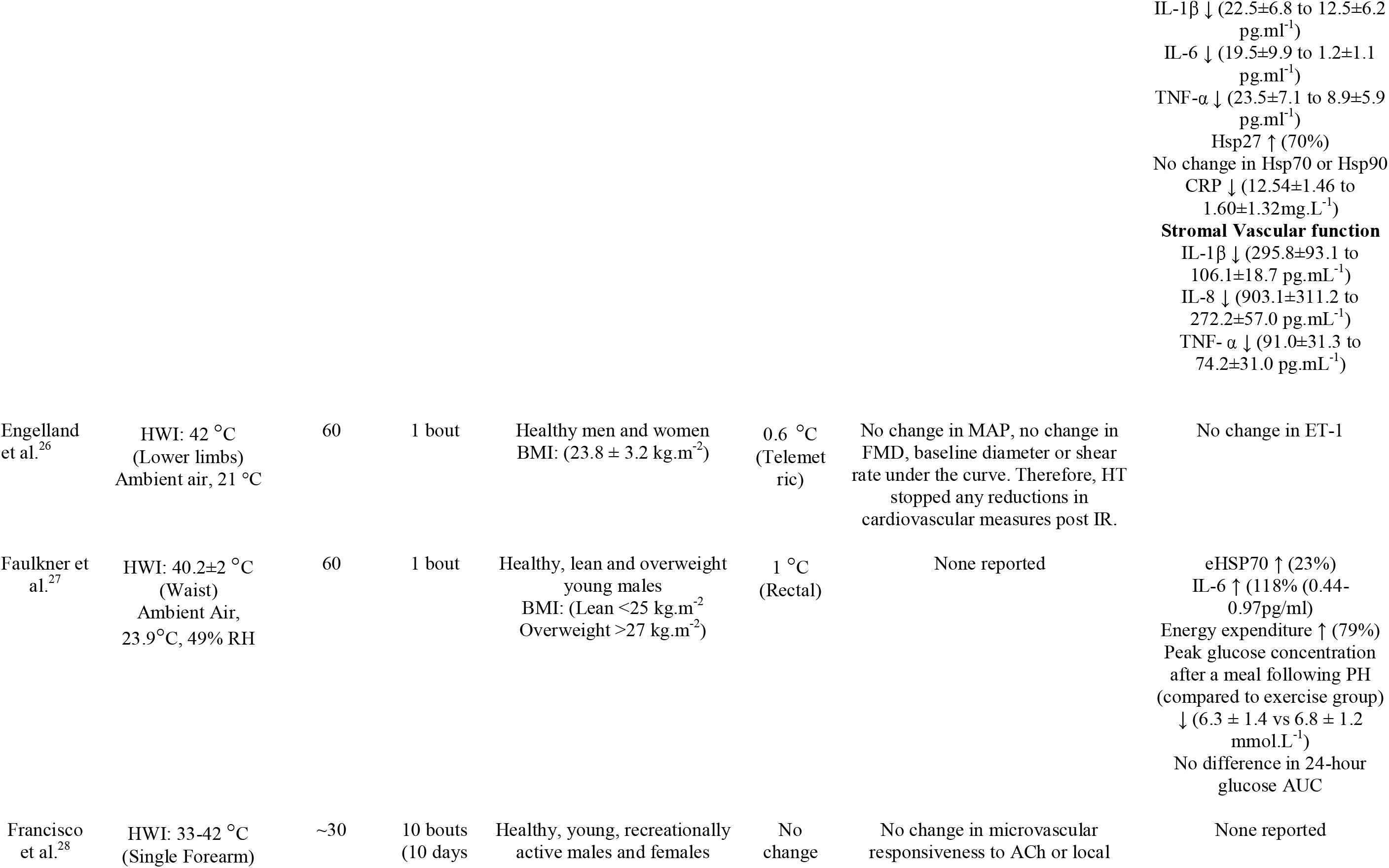
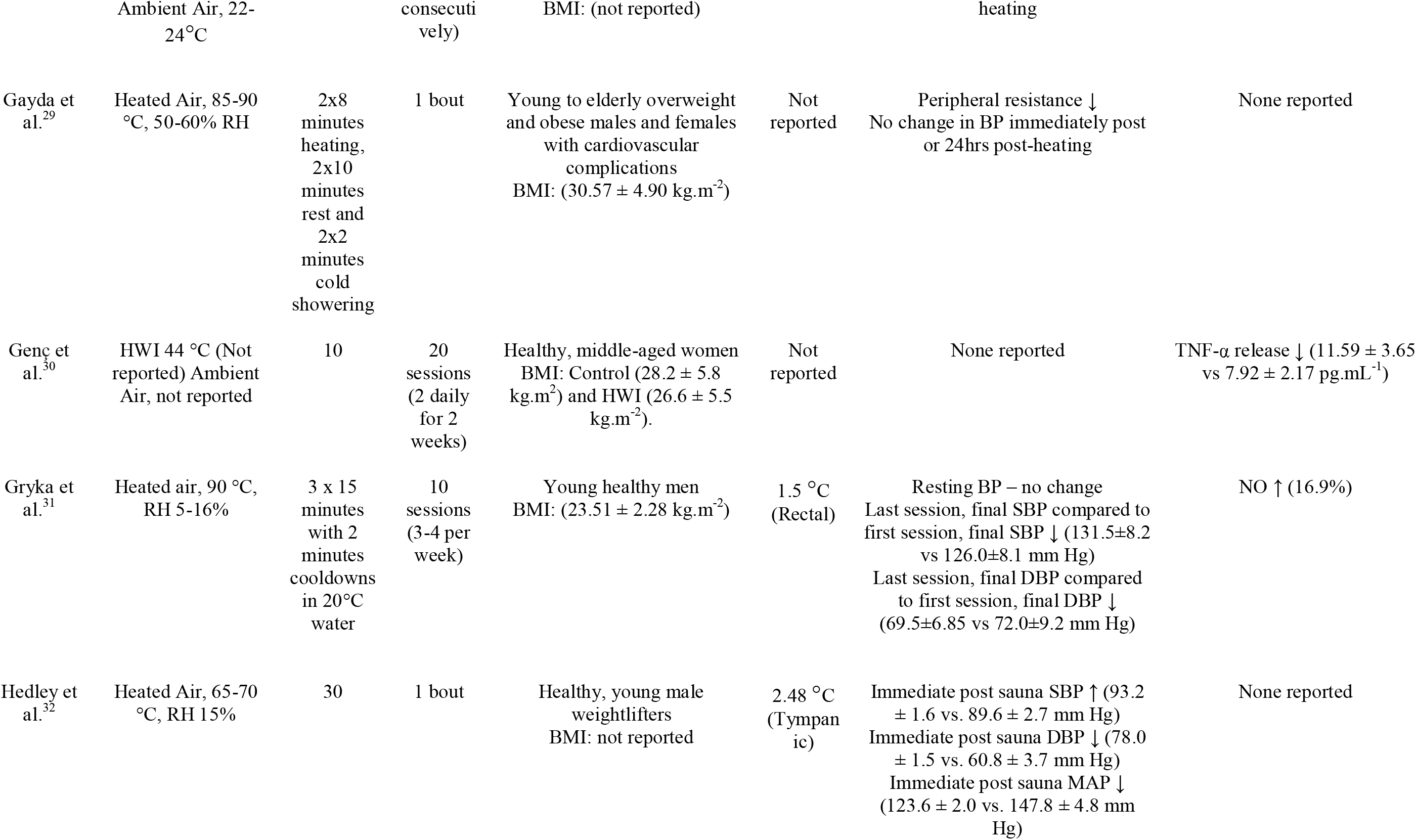
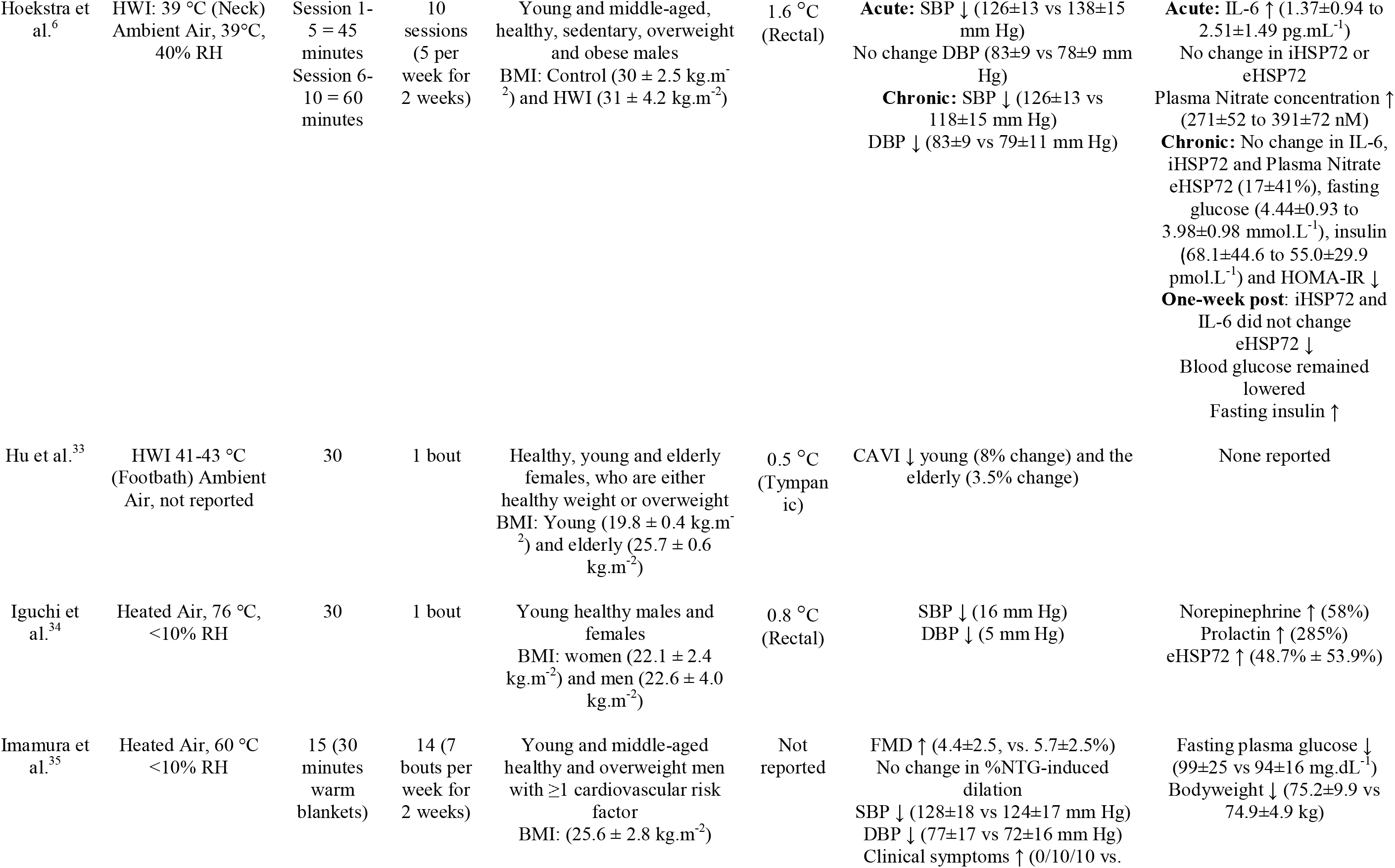
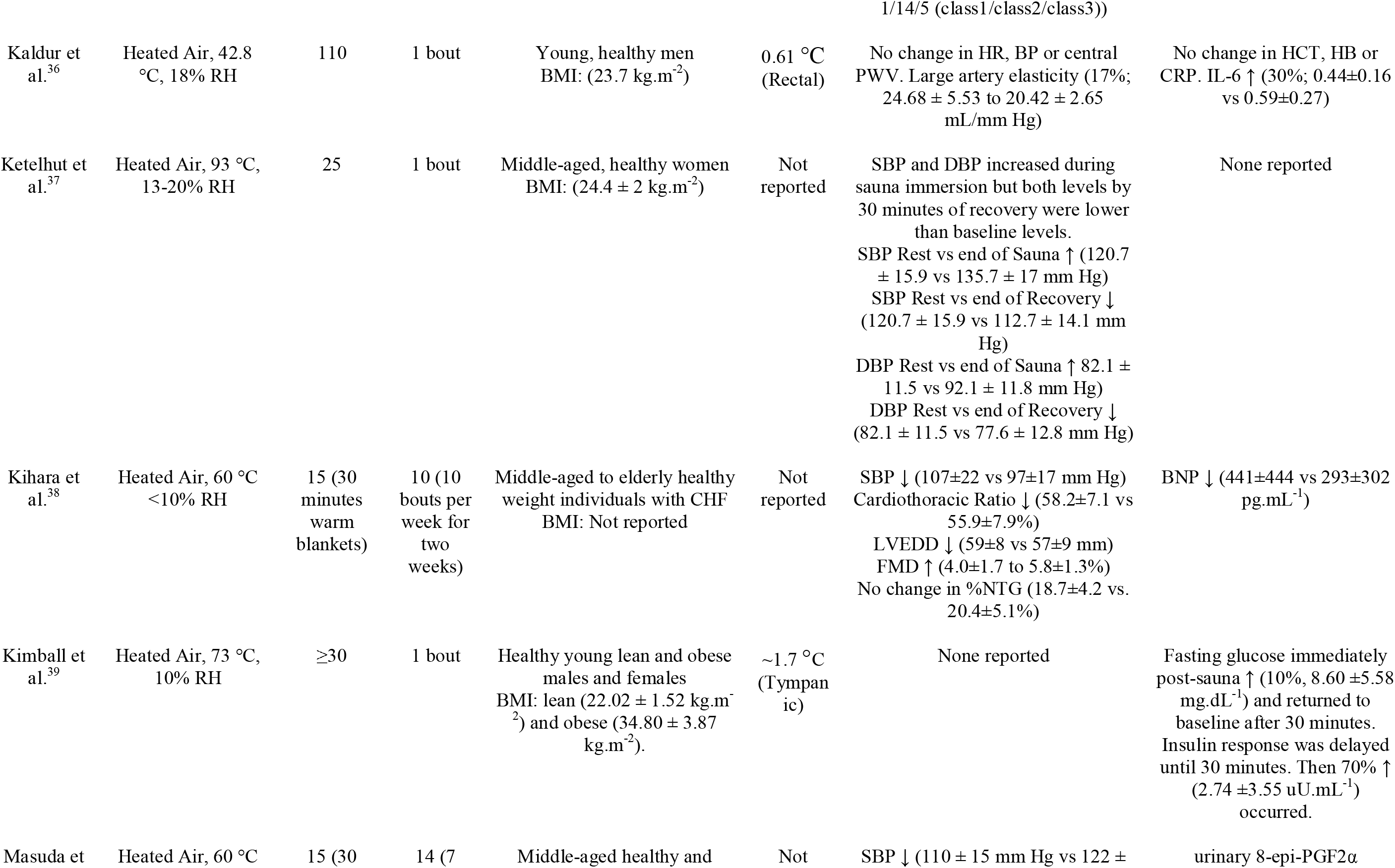
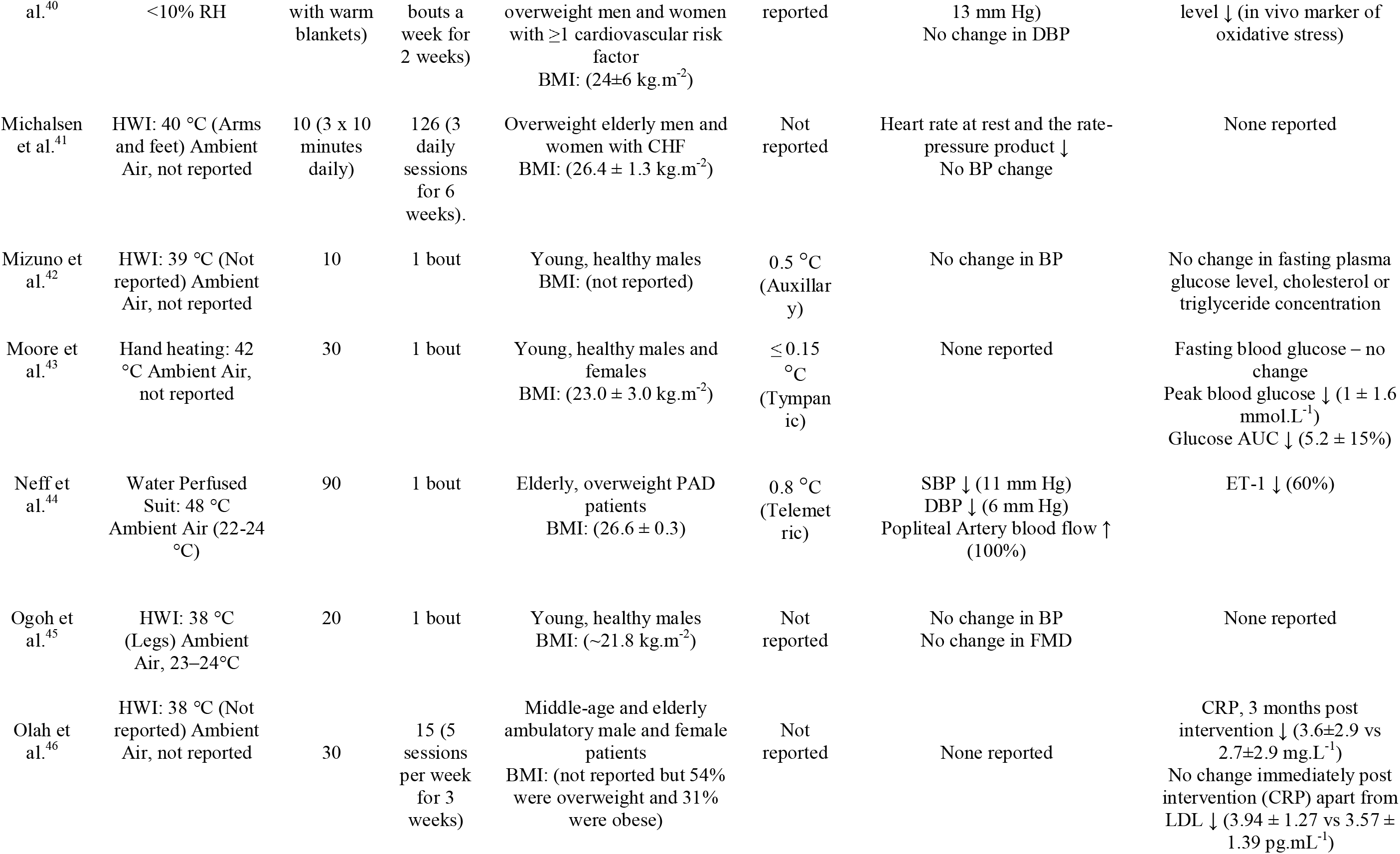
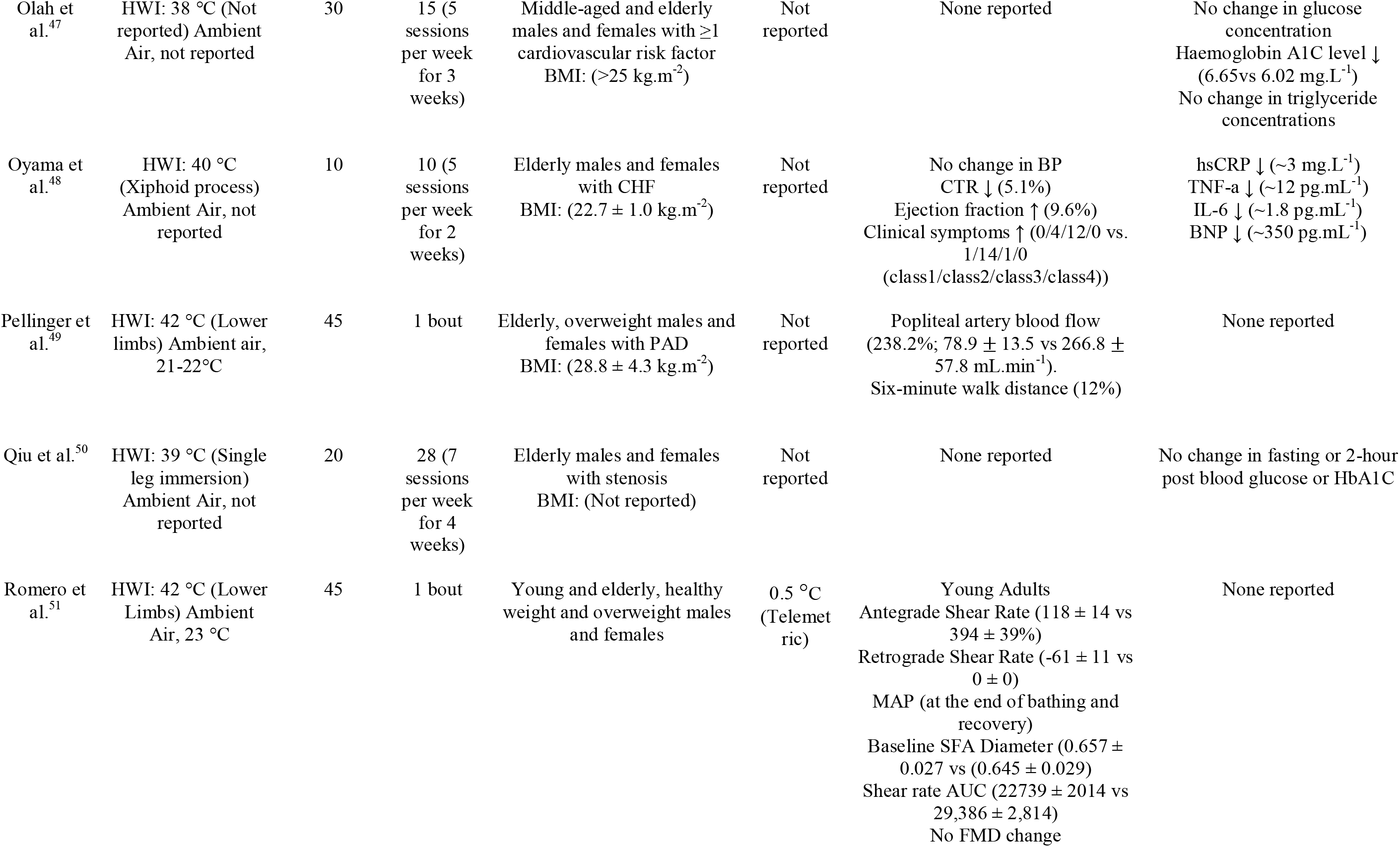
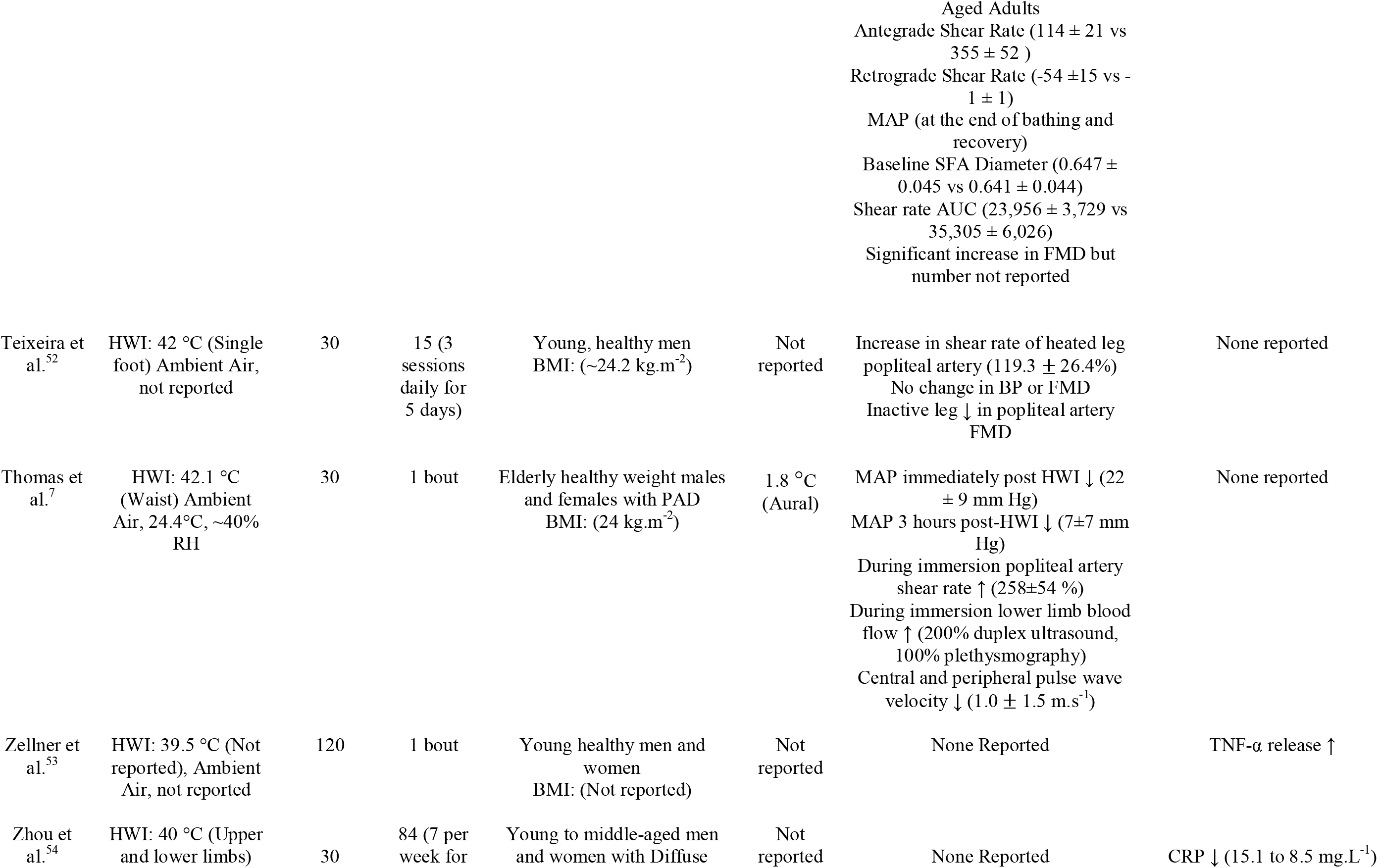
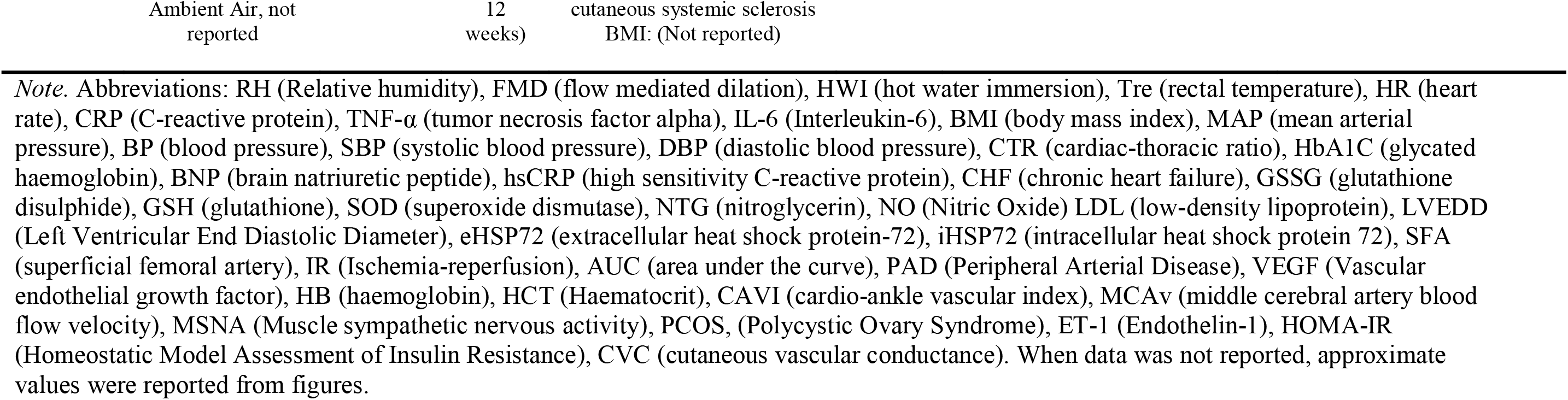
Heat Thermotherapy Intervention Papers Included in the Narrative Synthesis

**Table 3.** Change in key Cardiovascular and Cardiometabolic Measures following HT intervention

| Measures | Total no. of papers | Papers reporting beneficial health change | Papers reporting negative health change | Papers reporting no change | Change Values Reported | Average change | Range of change |
| --- | --- | --- | --- | --- | --- | --- | --- |
| <u>Cardiovascular</u> |  |  |  |  |  |  |  |
| FMD | 10 | 6 | 0 | 4 | 0, 0, 0, 0, 0, 1.3, 1.7, 1.8, 3.9, 5.3% | 1.6% | 0 to 5.3% |
| MAP | 13 | 5 | 0 | 8 | -23, -22, -13, -10, -5, 0, 0, 0, 0, 0, 0, 0 mm Hg | -5.6 mm Hg | 0 to -23 mm Hg |
| SBP | 11 | 10 | 1 | 0 | -16, -15, -11, -12, -10, -10, -8, -8, -5.5, -4, 3.6 mm Hg | -8 mm Hg | 3.6 to -16 mm Hg |
| DBP | 10 | 9 | 0 | 1 | -18, -11, -9, -6, -5, -5, -4, -5, -3.5, 0 mm Hg | -8.7 mm Hg | 0 to -18 mm Hg |
| SFA Compliance | 1 | 1 | 0 | 0 | 0.03 mm<sup>2</sup> mm.Hg<sup>-1</sup> | 0.03 mm<sup>2</sup> mm.Hg<sup>-1</sup> | 0.03 mm<sup>2</sup> mm.Hg<sup>-1</sup> |
| Pulse Wave Velocity | 5 | 3 | 0 | 2 | 0, 0, -72, -1, -1 m.s<sup>-1</sup> | -14.8 m.s<sup>-1</sup> | -72 to 0 m.s<sup>-1</sup> |
| Carotid intima media thickness | 2 | 2 | 0 | 0 | -0.06 mm | -0.06 mm | -0.06 mm |
| CAVI | 1 | 1 | 0 | 0 | 8% | 8% | 8% |
| CTr | 1 | 1 | 0 | 0 | -2.3% | -2.3% | -2.3% |
| CVC | 1 | 1 | 0 | 0 | 11% | 11% | 11% |
| Antegrade shear rate | 5 | 5 | 0 | 0 | 100, 119, 238, 258, 276% | 198% | 119 to 276% |
| MCAv | 1 | 1 | 0 | 0 | 2.3 cm.s<sup>-1</sup> | 2.3 cm.s<sup>-1</sup> | 2.3 cm.s<sup>-1</sup> |
| <u>Cardiometabolic</u> |  |  |  |  |  |  |  |
| IL-6 | 5 | 2 | 2 | 1 | -18.3, ~-1.8, 0, 0.54, 0.15 pg.ml<sup>-1</sup> | -3.9 pg.ml<sup>-1</sup> | -18.3 to 0.54 pg.ml<sup>-1</sup> |
| iHSP72 | 1 | 0 | 0 | 1 | 0 | 0 | 0 |
| eHSP72 | 2 | 1 | 1 | 0 | -17, 49% | 16% | -17 to 49% |
| Plasma nitrate | 1 | 1 | 0 | 0 | 120 nM | 120 nM | 120 nM |
| Glucose |  |  |  |  |  |  |  |
| a. immediately post-intervention | 2 | 0 | 2 | 0 | 0.5 mmol.L<sup>-1</sup> | 0.25 mmol.L<sup>-1</sup> | 0.5 mmol.L<sup>-1</sup> |
| b. fasting | 5 | 2 | 0 | 3 | -0.46, -0.3, 0, 0, 0 mmol.L<sup>-1</sup> | -0.15 mmol.L<sup>-1</sup> | 0 to -0.46 mmol.L<sup>-1</sup> |
| c. postprandial | 2 | 2 | 0 | 0 | -0.89, -0.5 mmol.L<sup>-1</sup> | -0.7 mmol.L<sup>-1</sup> | -0.89 to -0.5 mmol.L<sup>-1</sup> |
| Insulin |  |  |  |  |  |  |  |
| a. immediately post-intervention | 2 | 0 | 2 | 0 | 19 pmol.L<sup>-1</sup> | 9.5 pmol.L<sup>-1</sup> | 19 pmol.L<sup>-1</sup> |
| b. fasting | 2 | 1 | 0 | 1 | -13 pmol.L<sup>-1</sup> | -13 pmol.L<sup>-1</sup> | -13 pmol.L<sup>-1</sup> |
| Energy expenditure | 1 | 1 | 0 | 0 | 79% ↑ | 79% ↑ | 79% ↑ |
| Hb1Ac | 2 | 1 | 0 | 1 | 0, -0.63 mg.L<sup>-1</sup> | -0.32 mg.L<sup>-1</sup> | 0 to -0.32 mg.L<sup>-1</sup> |
| CRP | 4 | 4 | 0 | 0 | -10.94, -6.6, -3, -0.9 mg.L<sup>-1</sup> | -5.36 mg.L<sup>-1</sup> | -10.94 to -0.9 mg.L<sup>-1</sup> |
| BNP | 2 | 2 | 0 | 0 | ~-350, -148 pg.mL<sup>-1</sup> | 249 pg.mL<sup>-1</sup> | -350 to -148 pg.mL<sup>-1</sup> |
| TNF-α | 4 | 3 | 1 | 0 | -14.8, ~-12, -3.67 pg.mL<sup>-1</sup> | -5.2 pg.mL<sup>-1</sup> | -12 to -3.67 pg.mL<sup>-1</sup> |
| Triglycerides | 2 | 0 | 0 | 2 | n/a | n/a | n/a |

*Note.* Abbreviations: FMD (flow-mediated dilation), HWI (hot water immersion), Tre (rectal temperature), HR (heart rate), CRP (C-reactive protein), TNF-α (tumor necrosis factor-alpha), IL-6 (Interleukin-6), BMI (body mass index), MAP (mean arterial pressure), BP (blood pressure), SBP (systolic blood pressure), DBP (diastolic blood pressure), CTR (cardiac-thoracic ratio), HbA1C (glycated haemoglobin), BNP (brain natriuretic peptide), hsCRP (high sensitivity C-reactive protein), CHF (chronic heart failure), LVEDD (Left Ventricular End Diastolic Diameter), eHSP72 (extracellular heat shock protein-72), iHSP72 (intracellular heat shock protein 72), SFA (superficial femoral artery), CAVI (cardio-ankle vascular index), MCAv (middle cerebral artery blood flow velocity), CVC (cutaneous vascular conductance). When data was not reported, approximate values were reported from figures.

**Figure 3.**
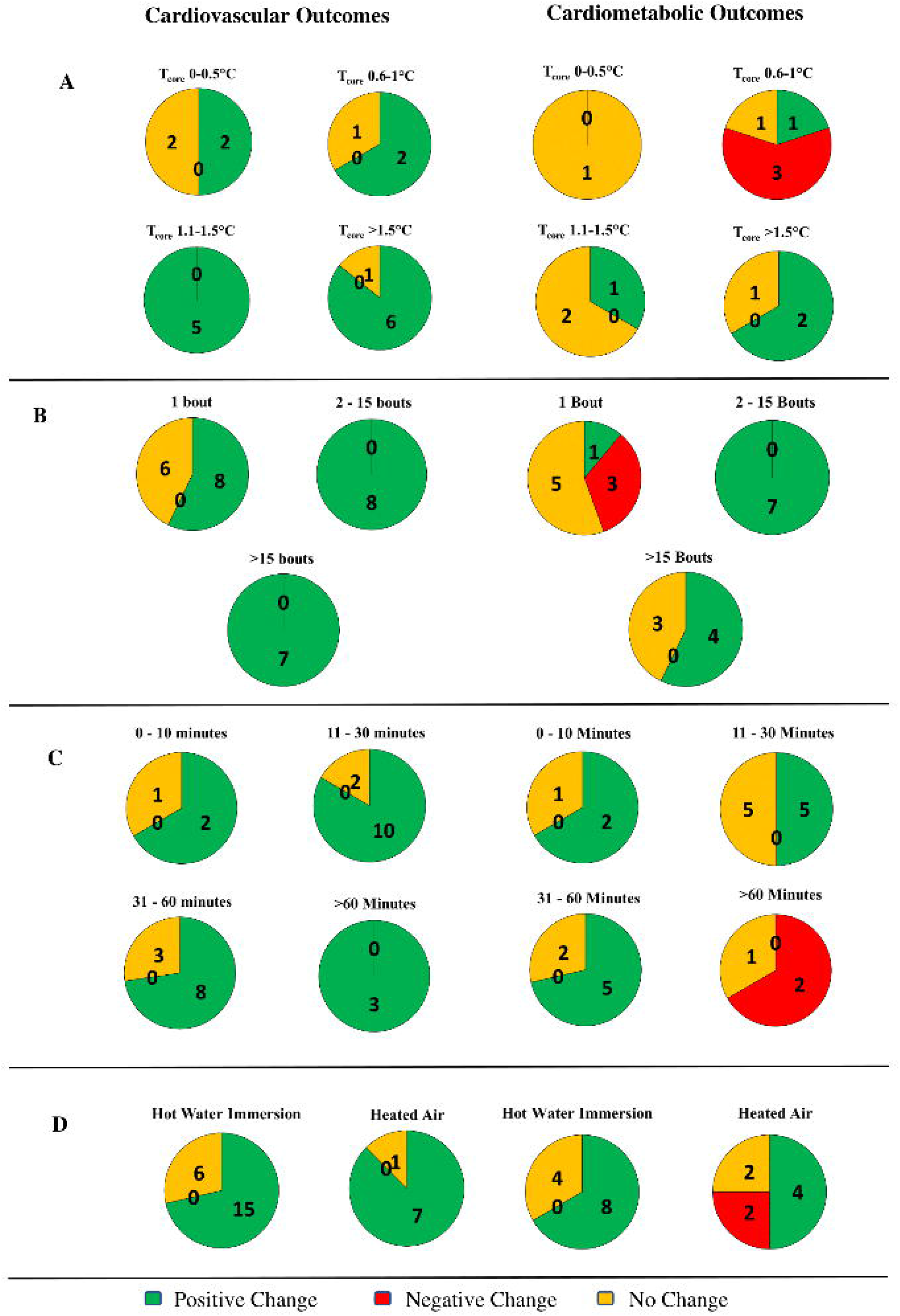

### Narrative Synthesis Robustness

A Risk of Bias assessment was completed for all papers in the Narrative Synthesis. In domain one, potential bias arising from the randomisation process, papers were deemed low risk (5/39) and some concerns (27/39). In domain two, potential bias due to deviations from intended interventions, papers were deemed no risk (38/39) and high risk (1/39). In domain three, potential bias due to missing outcome data, papers were deemed low risk (32/39) and some concerns (7/39). In domain four, potential bias in the measurement of the outcome, papers were deemed low risk (39/39). In domain five, potential bias in the selection of the reported result, papers were deemed low risk (2/39) and some concerns (37/39). For the overall risk of bias, papers were deemed some concerns (31/39) and high risk (8/39).

## DISCUSSION

### Main Findings

This is the first systematic review of the evidence from HT intervention studies reporting cardiovascular and cardiometabolic health outcomes. The main finding was that HT consistently improved cardiovascular and cardiometabolic health outcomes across different population groups (age, disease status, etc.), despite heterogeneous HT methodologies between studies. Specifically, over 74% of studies reported either a positive cardiovascular or cardiometabolic health benefit because of an HT intervention. These benefits included cardiovascular improvements in FMD, pulse wave velocity (PWV) and reductions in blood pressure. Cardiometabolic health improvements included reductions in inflammation and postprandial glucose concentration, and increased plasma nitrate concentration, all of which are associated with a reduction in overall CVD risk and mortality.^18^ Collectively, findings from this systematic review demonstrate the potential for HT to be a viable avenue to help prevent non-communicable disease progression in young sedentary populations, or attenuate cardiovascular or cardiometabolic disease progression (e.g. CVD or Type Two Diabetes Mellitus (T2DM)).

### Participant and Thermotherapy Method Characteristics

For the 39 included studies, participants ranged considerably in terms of their age and health status. Positive cardiovascular and cardiometabolic health outcomes in young healthy, albeit sedentary, adults are perhaps surprising. This is due to the assumption that in the young, endothelial functioning and arterial health is robust enough to prevent a decline or improvement in cardiovascular/cardiometabolic health.^55^ Positive cardiovascular and cardiometabolic health outcomes were also seen in older cohorts and cohorts with CVD complications. Overall, HT demonstrated positive cardiovascular and cardiometabolic health outcomes consistently across different population demographics despite varying HT type, durations, and temperatures.

HT type has an important role to play in the thermal load participants are exposed to and, potentially, the subsequent cardiovascular/cardiometabolic response. For HWI the most frequent exposure was hot water above 39.9 °C (see *Table 1*). The rationale for using a hotter, longer HT stimulus likely comes from heat acclimation literature, where sustaining a core body temperature above 38.5 °C is associated with increased proliferation of HSP and successful heat acclimation.^56 57^ Another reason may be that it is purported that antegrade shear rate patters in the limbs are greater when core body temperature is above 38.5 °C and sustaining this type and magnitude of shear rate results in greater cardiovascular adaptations.^11^ However, sustaining such high body/limb temperatures may not be obligatory, as several studies included herein reported positive cardiovascular and cardiometabolic health benefits despite maintaining core body temperature below 38.5 °C.^27 33 34^

Heated air exposure, typically via sauna or steam room visits, are a common form of HT, especially in Scandinavian countries e.g. Finland.^12^ For the included studies that used heated air exposure, HT interventions typically lasted 16 to 32 minutes, which resulted in an average body core temperature increase of 0.8 to 1.7°C, compared to 1.5 to 1.6°C in HWI interventions. Despite the difference in core temperature change, both interventions showed improvements in cardiovascular and cardiometabolic outcomes. Unfortunately, it is difficult to assess whether core temperature increase is the cause of cardiovascular or cardiometabolic adaptations, as 18/39 papers did not report the change in body core temperature induced by the HT exposure, and the 21/39 studies measuring core temperature used different body sites for temperature measurement. Future studies should consider using valid and reliable core temperature measures to investigate the relationship between core temperature change and the subsequent cardiovascular/cardiometabolic response.

To ensure a cardiovascular response to HT, immersion to the shoulder/neck may be needed to ensure an adequate thermal load. For example, frequent ten-minute partial body HWI resulted in no cardiovascular and cardiometabolic changes.^41^. In terms of sauna bathing, 15 minutes of hot air exposure followed by 30 minutes of blankets in ambient environmental conditions resulted in cardiovascular and cardiometabolic improvements with participants of varying age and health status.^35 38^ Again, these papers used a short duration protocol, indicating that it is possible to have beneficial outcomes with a short exposure to HT if there are enough bouts, a strong enough thermal stimulus, and enough body surface area exposed. These short durations also indicate that there does not need to be an increase of over 1.5-2 °C in rectal core temperature for positive cardiovascular and cardiometabolic outcomes, although most short duration papers in the narrative synthesis did not report any core temperature recordings to confirm this.

### Measurements of Cardiometabolic and Cardiovascular Health

The most consistent measurement of cardiovascular health was FMD, with 6/10 papers reporting a 1.3 – 5.3% positive change following HT. The remaining four studies reported no significant change in FMD following HT, although two studies found FMD decreases in control groups following five days of physical inactivity or 40 minutes of forearm ischemia-reperfusion.^23 52^ Thus, HT appears to improve and even protect cardiovascular function against insults such as physical inactivity and ischemia-reperfusion injury. The most frequently reported cardiovascular outcome was blood pressure (Mean Arterial Pressure (MAP), Systolic Blood Pressure (SBP) and Diastolic Blood Pressure (DBP) respectively)). On average, SBP, DBP and MAP reduced by −6 to −9 mmHg, which is clinically meaningful since population wide reductions in SBP of modest magnitude (2 mm Hg) are predicted to have a substantial impact on CVD prevention.^58^ The most consistent measurement for a positive cardiometabolic outcome was the inflammation marker CRP, with 4/4 studies reporting a significant reduction following the HT intervention. CRP is a surrogate marker for insulin resistance and is strongly associated with atherosclerotic disease.^59^ Therefore, reductions in CRP and other inflammatory markers as shown in *Table 5* indicate cardiometabolic health benefits when exposed to HT.

Although it may be a strength that different studies reported different, positive cardiovascular and cardiometabolic outcomes across a range of cohorts, it also makes it difficult to state whether these specific outcomes are consistent and reproducible. Moreover, it is important to understand the relationship of how one health outcome compares with other outcome measurements within the same participants. For example, alongside an increase in brachial FMD, an indicator of improved endothelial function and potentially cardiovascular health, other measurements such as pulse wave velocity, MAP and Cardiovascular Conductance (CVC) would provide a more conclusive examination of cardiovascular health. Indeed, this was the case for cardiovascular measures in young sedentary participants^21 22^ and elderly participants^7^ reviewed herein, but our analysis revealed that a consistent battery of cardiovascular assessments has not been conducted in a middle-aged overweight cohort. This is surprising because middle-aged overweight/obese cohorts are at high-risk for insulin resistance and non-communicable disease progression, which typically is associated with cardiovascular health complications.^18^ Regarding cardiometabolic health, a battery of assessments has been conducted in overweight, middle-aged participants,^6 27^ but not in elderly populations or cohorts that are not obese but have cardiovascular risk factors or congestive heart failure; populations that are also at high risk of developing metabolic syndrome.^18^ To understand the cardiovascular and cardiometabolic health outcomes across different population cohorts and to better prescribe tailored HT health interventions, especially for those who are at more risk of CVD complications, studies need to incorporate these test batteries into future research trials.

### Participant Health Status, Age and Thermotherapy Outcomes

Of studies that included overweight/obese individuals, 9/10 studies reported positive cardiovascular outcomes and 9/10 studies reported beneficial cardiometabolic outcomes following an HT intervention. One study, however, reported a negative cardiometabolic outcome (i.e. increased fasting glucose concentration and delayed insulin response) after a one-off, single bout of HT.^60^ Similarly, in young, healthy men and women, one bout of HT resulted in immediate increases in fasting glucose, insulin and TNF-α.^61 62^ It is unclear why an acute bout of HT appears to impair glucose control, particularly given the positive effects of chronic HT (10-14 bouts) on cardiometabolic outcomes.^6 35^ A possible reason may be the residual effect of stress hormones, which are elevated after HWI.^63^ These stress hormones can temporarily impair insulin sensitivity and increase hepatic glucose output.^64^

Of studies that included healthy individuals (e.g., not obese, no CVD etc.), 11/16 studies reported beneficial cardiovascular outcomes including reduced blood pressure, carotid media thickness and increased FMD.^22^ Notably, three of the studies that did not report improved cardiovascular outcomes following HT did prevent a reduction in FMD in the brachial^23 26^ and femoral^52^ arteries relative to a control group, where a reduction in FMD occurred across the control period. In contrast, cardiometabolic outcomes following HT were more inconsistent in healthy cohorts, with 5/10 of studies reporting positive outcomes, 4/10 studies reporting negative outcomes,^60 61^ and one study reporting no change following an HT intervention.^65^ Given that healthy individuals typically are not insulin resistant nor have increased levels of systemic inflammation, which are factors associated with cardiometabolic dysfunction,^18^ such findings are perhaps unsurprising. Alternatively, the thermal load used in these studies^42 45^ may not have been sufficiently large enough to evoke a cardiometabolic response. For example, these studies used protocols of brief duration^42^ (10 minutes) or partial limb heating protocols.^45^ Interestingly, studies that reported negative cardiometabolic outcomes also had significantly longer HT durations than other HT interventions (120-240 minutes at 33°C air temperature, 44% RH;^24^ vs narrative synthesis average of 53 minutes; *Table 4*). These prolonged exposures may have negative effects on those who can normally regulate serum glucose concentrations^24^ by promoting a prolonged, pro-inflammatory response as shown by the increase in TNF-α.^45^

All elderly participants were either overweight/obese or possessed a cardiovascular risk factor. Within this elderly cohort, 6/8 studies reported a positive cardiovascular outcome and 4/5 studies reported a positive cardiometabolic outcome. Cardiovascular improvements may be more apparent in elderly participants in comparison to younger cohorts due to age-related changes in vasculature compliance, endothelial function and underlying increases in atherosclerotic plaque.^66^ Cardiometabolic improvements may also be more apparent as HT could attenuate age-related increases in the chronic inflammatory state caused by cell senescence and innate immunity dysregulation.^67^

## LIMITATIONS

### Risk of Bias Assessment

No studies selected within the narrative synthesis achieved overall a low-risk score. In the domain bias, included articles were classified as “high risk” because they purposely matched participants by age and body mass, removing any randomisation process. Given the experimental nature and therefore smaller *n* used in these studies, appropriate matching of covariates is necessary to examine the hypothesis. It highlights the need, however, for randomised control studies to be conducted to progress the use of HT as an alternative treatment. Other included studies were classified as “some concerns” because, despite reporting random sequencing, they did not state how this occurred (e.g. computer sequencing). Another area of concern was the “risk of bias in the selection of the reported result” domain (see *Table 7*), where most studies were marked “some concerns”. This was due to papers not reporting a pre-specified plan of how the results will be reported. The overall level of risk for the included studies reflects the stage of HT research, where studies are still mechanistic in nature. To summarise, despite strong methodological processes and most papers using non-subjective measurements, there are some concerns of a type one or two error risk for papers included in the narrative synthesis.

### Systematic Review Limitations

The main strength of this systematic review was the ability to select a wide range of studies using different methodologies and outcome measures. This enabled us to assess the overall question of whether HT improves cardiovascular and cardiometabolic health across different participant profiles and HT methodology. However, due to the broad range of population groups and measures used to assess cardiovascular and cardiometabolic function, it is difficult to compare the strengths and weaknesses of different HT interventions and to determine which HT method produces the most optimal cardiovascular/cardiometabolic outcome. Subsequently, a meta-analysis could not be performed at this time. To generate the conditions needed to conduct a metanalysis and to empirically assess heat thermotherapy strategies, more papers are needed that use a consistent participant profile, cardiovascular/cardiometabolic health outcome measures and HT methodology.

### Overall Narrative Synthesis Robustness

A strength of this narrative synthesis is that all included papers were controlled trials using reliable and objective measures. Included studies were selected using a systematic approach, thus minimising the risk of researcher bias. To test the robustness of the narrative synthesis, a risk of bias assessment was performed that concluded there was some level of risk with the papers included. Due to these processes, we feel confident that the results of the narrative synthesis are a true reflection of HT research to date, and the low percentage of high-risk papers was deemed insufficient to affect the interpretation of the results from the review.

## FUTURE DIRECTIONS AND IMPLICATIONS

HT results in positive cardiovascular and cardiometabolic outcomes in a range of participant cohorts. However, the research to date has exclusively been experimental, mechanistic studies. Future research needs to move towards larger randomised control trials and address the issues raised in the Risk of Bias Assessment. Current evidence on HT decay is limited, with only two studies reporting cardiometabolic health outcomes post HT and no studies assessing the decay effect of cardiovascular outcomes. Therefore, information on how long these cardiovascular and cardiometabolic health benefits last for, and whether the decay effect is attenuated by the number of bouts (i.e. short-term vs a chronic HT intervention) is limited. It is also not known whether the decay effect can be attenuated by “top-up” bouts and the duration of intervention needed before the “top-up” bout is effective. Another factor to consider is adherence and enjoyment. Future HT interventions must assess psychophysical responses as well as adherence in study populations, to determine if HT is a viable alternative to more traditional lifestyle interventions (e.g. diet and exercise).

## CONCLUSION

The aim for this systematic review was to objectively assess whether HT improves cardiovascular and cardiometabolic health; by collating and analysing all causational designed HT papers with a cardiovascular or cardiometabolic outcome in a non-biased, reproducible manner. Despite a wide range of methodology, participant profiles and some concerns regarding the Risk of Bias Assessment, over 74% of papers reported positive cardiovascular/cardiometabolic health outcomes after an HT intervention. These findings indicate that HT is a promising therapy to protect and mitigate cardiovascular and cardiometabolic disease progression. Given the potential wide-ranging benefits of HT, further clinical research is warranted.

#### What is already known

- Treatments of cardiovascular disease (exercise and pharmaceutical interventions) can have limited impact and poor adherence rates.
- Heat Thermotherapy (HT) can address cardiovascular and cardiometabolic disease risk factors and has greater adherence rates than traditional treatments.
- No study to date has systematically reviewed the efficacy of HT to improve cardiovascular and cardiometabolic health outcomes.

#### What are the new findings

- A systematic review of the literature demonstrated that over 74% of included studies reported a positive cardiovascular or cardiometabolic health outcome following HT.
- HT improved cardiovascular and cardiometabolic health outcomes across a wide range of participant cohorts (age and health status), despite the various HT methodology used (duration of HT, number of bouts etc.).
- HT is a promising alternative therapy to reduce cardiovascular or cardiometabolic disease risk/progression in young sedentary populations as well as clinical populations.

## Data Availability

Data are available upon request at School of Sport, Exercise and Rehabilitation Sciences, University of Birmingham, UK.

## Acknolegements

We would like to thank Dr Andy Soundy and Ms Bethany Skinner for their assistance.

## Notes

### Competing Interest Statement

The authors have declared no competing interest.

### Funding Statement

No funding was obtained for this paper and no authors or their institutions at any time received payment or services from a third party for any aspect of the submitted work.

### Author Declarations

Approval is from the University of Birmingham.

